# A juxtaposition of safety outcomes between various doses of sodium-glucose co-transporter inhibitors, in insulin-treated type-1 diabetes mellitus patients: a protocol for systematic review and meta-analysis of double-blinded randomized controlled trials

**DOI:** 10.1101/2020.06.22.20137349

**Authors:** Sumanta Saha

**Affiliations:** None

**Keywords:** Diabetes Mellitus, Type 1, sodium-glucose cotransporter-2 inhibitor, insulin, side effect, Systematic review, RCT

## Abstract

**Aims:** Several clinical trials have tested the safety profile of sodium-glucose co-transport inhibitors’ (SGLTis) in adult type 1 diabetes mellitus (T1DM) patients. However, no systematic review has compared its variation between large and low dose SGLTis. Henceforth, a review protocol is proposed here to review it. Besides, it will compare the side effects of each of these interventions with the placebo.

**Methods:** Different electronic databases will be searched for randomized double-blinded placebo-controlled trials (published in the English language) studying the above objective, irrespective of their publication date. After selecting the eligible trials, their data on the study design, population characteristics, compared interventions, and outcomes of interest will be extracted. Then, utilizing the Cochrane tool, each trial’s risk of selection bias, detection bias, performance bias, attrition bias, reporting bias, and other bias will be judged. Next, depending on clinical heterogeneity among the trials, a random-effect or fixed-effect model meta-analysis will be used to compare the respective outcomes. Via the Chi^2^ and I^2^ statistics, the statistical inconsistency among the trials will be estimated. When this is substantial, subgroup analysis will follow. Publication bias will be evaluated by funnel plots and Egger’s test. A sensitivity analysis will be done to check different assumptions. If a quantitative juxtaposition is not possible, a narrative reporting will ensue. Conclusions: The proposed review will compare the safety profile between the mega and low dose SGLTis in insulin-treated T1DM patients. Besides, each of these two types of doses will be compared with placebo for the same.

**REGISTRATION:** PROSPERO (Registration no. CRD42019146578)

## INTRODUCTION

The autoimmune destruction of insulin (an anabolic hormone) producing pancreatic beta-cells leads to type 1 diabetes mellitus (T1DM), a disease characterized by hyperglycemia.(1,2) Although the exact etiology of the disease is unknown, the genetic predisposition may have some role. Individuals with certain human leukocyte antigen alleles (DR and DQ) are at greater risk of developing T1DM upon exposure to various trigger agents like viruses, environmental toxins, and dietary factors.(1) T1DM frequently begins between 4-6 and 10-14-year-olds, although it can start at any age.(1) About 5-10% of diabetes patients are T1DM patients.(1) Complications of T1DM often include neuropathy, nephropathy, retinopathy, hypoglycemia, DKA, cardiomyopathy, and diabetic foot disease.(1) To prevent these hyperglycemic complications in T1DM patients, insulin, the mainstay of treatment, is required lifelong.(1–3)

However, this entire insulin dependence doesn’t suit every T1DM patient, because of the inconveniences of insulin therapy like multiple insulin injections and finger pricks every day to control and self-monitor the blood glucose levels, respectively.(4,5) Besides, there are adverse consequences of intensive insulin therapy on the health like lipohypertrophy or atrophy (when insulin is repeatedly injected at the same site), unwanted weight gain, raised risk of hypoglycemic episodes, and glycemic variations.(6) Some of these complications of absolute insulin therapy might affect the health and treatment of T1DM patients further. For instance, the subsequent injections in insulin-induced lipohypertrophied areas can slow down insulin absorption.(6) Likewise, the insulin-induced weight gain might raise the risk of cardiometabolic complications by decreasing compliance with the insulin regimen.(6) Therefore, to reduce the discomforts and complications of intensive insulin therapy, the role of insulin-independent adjunct therapeutics are vital in the treatment of T1DM patients. In this regard, sodium-glucose co-transport inhibitors (SGLTi) have drawn significant attention of the medical community.

SGLTis are phlorizin compounds.(2,4) By inhibiting SGLT1 and SGLT2 receptors, SGLTis cause glycosuria and decreased intestinal absorption of glucose.(4) SGLT2 receptors in the proximal convoluted tubule reabsorb almost 90% of the filtered glucose.(6–8) Examples of SGLT2 inhibitors include dapagliflozin, empagliflozin, and canagliflozin.(9) SGLT1 is predominantly found in the intestine, and its inhibition regulates the blood glucose level by increasing the release of gastrointestinal hormones through increased glucose delivery to the distal small intestine.(6) Sotagliflozin is a dual SGLT2/1 blocker; its capacity to inhibit SGLT1 receptors provides an additional benefit in decreasing the glucose levels postprandially.(2)

Existing early stage trials suggested several efficacies of SGLTi in T1DM patients. Such trials comparing SGLT2 inhibitors (dapagliflozin: 5 mg and 10 mg, empagliflozin: 25 mg, 10 mg, and 2.5 mg, and canagliflozin: 100 mg and 300 mg) to placebo in insulin-treated inadequately controlled T1DM patients, demonstrated an improvement in glycemic control, body weight, and insulin requirement.(10–12) Likewise, trials that investigated sotagliflozin in T1DM patients and type 2 diabetes mellitus (T2DM) patients, observed improvement in glycemic control (by limiting the post-meal glycemic excursions), weight control, and daily insulin requirements.(13) A recent meta-analysis suggested that mega-dose empagliflozin treatment in insulin-treated T1DM patients with optimum renal functioning aids in achieving better glycemic control, compared to the placebo.(14)

However, despite these benefits, due to safety concerns, both sotagliflozin(15) and SGLT2is are currently not approved by the U.S. Food and Drug Administration.(16) Therefore, it is crucial to research the safety of SGLT extensively. Contemporarily, several randomized controlled clinical trials have reported the safety profile of SGLT inhibitors in T1DM patients.(12,17–19) Although a statistical comparison of these side effects between the treatment arms was not available, based on its frequency, some idea about the safety profile of SGLTi can be made. The trials(12,17,18) on T1DM patients that compared the safety profile between large and low dose empagliflozin, canagliflozin, dapagliflozin observed that the overall percentage of side-effects due to any cause were higher in the recipients of the former. In these large dose recipients this was 100%, 68%, and 67% in empagliflozin (25 mg), canagliflozin (300 mg), and dapagliflozin (10 mg), respectively.(12,17,18) While the serious side effects did not happen with any dosages of empagliflozin in a trial,(17) in dapagliflozin recipients, it was more frequent with the low dose (5mg, 7%) than the large dose (10mg, 2.6%).(18) The serious side effects were found in 12.5% of T1DM patients using 400 mg sotagliflozin in a trial.(19) In T1DM patients, the proportion of hypoglycemic side effects was dose irrespectively high in all canagliflozin users (98-99%),(12) but in empagliflozin users, it varied dose-wise (2.5mg: 84%, 10mg: 68%, 25mg: 94%).(17)

Given the safety concerns leading to non-approval of these drugs’ use in T1DM patients by the US FDA and agglomeration of available clinical trials that tested their safety profile, it is imperative to synthesize novel evidence in this regard. Interestingly, the existing review articles have primarily compared the safety profile of SGLTi with the placebo.(20–23) However, no systematic review has yet compared the side effects profile between large and low doses of SGLTis. Therefore, to explore this under-reviewed area, this systematic review and meta-analysis protocol aims to study this comparison. Additionally, with large and low dose SGLTi recipients, it will also compare these outcomes to placebo recipients, respectively.

## METHODS

The proposed review’s inclusion criteria will be the following: 1. Study design: Double-blinded, parallel-arm (any number of arms) randomized placebo-controlled trials. 2. Participants: Adult (18 years or older) insulin-treated T1DM patients irrespective of their gender. The diagnosis of T1DM will be accepted as per the trialists. 3. Intervention arm: The intervention groups should receive a mega-dose of dapagliflozin (10 mg), empagliflozin (25 mg), canagliflozin (300 mg), or sotagliflozin (400 mg) every day. The determination of these mega doses was based on the maximum dose in which they were tested in T1DM patients in different clinical trials.(12,17–19) 4. Comparator arm: The comparator group/s should receive the same drugs as the intervention arm, however, at a lower dose. Comparator arm receiving placebo will be also recruited. 5. Outcome: The following will be the primary outcomes of interest-overall side effects due to any cause, attrition due to adverse effects, side effects due to the tested intervention, and serious adverse effects.

The secondary outcomes of interest will include the number of participants experiencing diabetic ketoacidosis, hypoglycemia, genital infection, and urinary tract infection. To be included in the proposed review, the trials must report at least one of the primary outcomes. The secondary outcomes will not form the inclusion criteria.

All outcomes should be reported in participants who have received at least one dose of the test medication or placebo after randomization. The definitions of these outcomes will be accepted as per the trialists.

The exclusion criteria incorporate the following: 1. Trials in which the trial population is diagnosed with type-2 diabetes, gestational diabetes, or maturity-onset diabetes of young. 2. Studies of other designs like crossover trials or quasi-experimental studies.

The proposed review protocol has been registered at the PROSPERO (registration no. CRD42019146578).(24) This protocol follows the PRISMA-P checklist.(25)

### Search for eligible trials

Next, the eligible trials’ titles and abstracts will be searched in various electronic databases (PubMed, Embase, and Scopus). The search will not be limited to any date, but it will be restricted to the English language only.

The search strategy to be used in the PubMed database is described here. Following search terms will be used: ‘safety’ OR ‘tolerance’ OR ‘adverse event’ OR ‘side effect’ OR ‘side-effect’ AND ‘canagliflozin’ OR ‘dapagliflozin’ OR ‘empagliflozin’ OR ‘sotagliflozin’ OR ‘sodium-glucose cotransporter’ OR cotransporter* OR sglt* AND ‘type-1’ OR ‘type 1’ AND ‘diabetes.’ The search will be narrowed down to clinical trials by using the filter ‘Randomized Controlled Trial’ and ‘Clinical Trial.’

For other databases without such filters, instead, the following search terms will be used - ‘trial’ OR ‘randomised’ OR ‘randomized’ OR ‘controlled.’

Additionally, eligible trials will also be searched in the references of papers included in the proposed review.

### Selection of eligible trials

The database search results will be uploaded to the Rayyan(26) systematic reviews software. Then, after excluding the duplicates, the titles and abstracts of papers will be read to find trials matching the above-mentioned eligibility criteria.

A paper will be read in entirety if it seems to be eligible for inclusion in this review or when a decision of inclusion or exclusion can’t be made by reading the excerpts alone. The list of publications excluded after reading full-text will be retained with their reasons for elimination. The entire study selection process will adhere to the Prisma 2009 flow diagram.(27)

When multiple trials source data from the same study population, one that counted the overall side effects (cause irrespective) based on the maximum number of side effects will be reviewed. If the tally persists, the trial with the longest follow-up time for adverse effects will be included in the review.

### Data extraction

From the recruited trials, data of study detail, participant characteristics, interventions compared, and outcomes of interest will be extracted. About study detail, the trial’s registration number, randomization method, blinding (single or double-blinded), duration, number of intervention arms, site (single centered or multicentric), participant consent, ethical clearance, country (where conducted), and funding information will be collected.

The following participant characteristics will be gathered - diagnosis, their number in each intervention arm, mean age, the average duration for which they have been diagnosed with T1DM, and their number for whom the outcome data of interest is not available. Concerning intervention information, the dosage and treatment regimen of each of the intervention arm will be collected. Finally, for the outcomes of interest, the number of participants experiencing it after taking at least one dose of the tested intervention will be extracted from each of the intervention groups.

### Risk of bias assessment

For each trial, using the Cochrane tool, the risk of selection bias, performance bias, detection bias, attrition bias, reporting bias, and miscellaneous bias will be evaluated and categorized as low, high risk, or unclear (if it does not meet the low or high-risk categorization).(28)

The selection bias will be assessed by the random sequence generation method used to allocate interventions to the participants and the means used to conceal this allocation from the participants. The performance bias will be based on the adequate blinding of study participants and personnel. Depending on the blinding of the outcome assessors, the detection bias will be judged. Based on the missing outcome data in each treatment arm, attrition bias will be evaluated. By comparing the variation in a trial’s reported findings from its protocol or pre-stated plans, the reporting bias will be assessed.

### Author roles

SS1 will perform the database search, upload the results to the systematic review software, and exclusion of duplicate papers. Then, the authors will independently select eligible trials, extract trial data, and critically appraise the studies. Any conflict in opinion among the authors will be settled by a discourse, and if it remains unresolved, a third-party opinion will be sought.

### Meta-analysis

Due to the dichotomous nature of the outcome data, for each outcome, risk ratios (RR) will be determined by a meta-analysis. It will compare between large and low dose SGLTis and each of these with placebo, respectively. In trials with more than one intervention arms testing the low dosages, the respective outcome data will be combined across these treatment arms. The summary estimate from meta-analysis will be analyzed using either a random-effect model (DerSimonian and Laird method) or a fixed-effect model (inverse variance method). This model choice will depend on the clinical heterogeneity of the trials like trial settings, study design, participant characteristics, etc. and not the pre-determined statistical inconsistency. If the reviewed trials are clinically heterogeneous, a random-effect model will be used or vice-versa.

If an outcome doesn’t occur in any of the contrasted intervention arms of a trial, it will not be included in the meta-analysis. However, when it happens in either of these arms, 0.5 will be added to each cell of the 2×2 table, and the trial will be incorporated in the meta-analysis.

For summary estimates of meta-analysis, a statistical significance will be determined at a p-value <0.05 (and 95% confidence interval).

The trials with a high risk of bias will not be included in the meta-analysis. Any outcome for which a quantitative juxtaposition is not possible, a narrative reporting will ensue.

### Heterogeneity and publication bias

Using the I^2^ and Chi^2^ statistics, the statistical inconsistency among the trials will be determined. At the I^2^ statistics values of 0-40%, 30-60%, 50-90%, and 75-100%, the heterogeneity will be categorized as less, moderate, substantial, and considerable, respectively.(28) At a p<0.1, the statistical significance of the Chi^2^ statistics will be estimated.(28)

If substantial or considerable statistical heterogeneity is detected in a meta-analysis of at least 10 trials, a subgroup analysis will be done. It will be done between the trials testing SGLT2 inhibitors and dual SGLT2/1 inhibitor, between the trials in which the participants’ estimated glomerular filtration rate (GFR) was less than 60 mL/min/1.73 m^2^ and more than 60 mL/min/1.73 m^2^, and based on missing outcome data.

Publication bias will be assessed using funnel plots. Additionally, an Egger’s test will be used when at least ten trials are available for meta-analysis.

### Additional analysis

The following types of sensitivity analysis will be done by repeating the meta-analysis-1. By using a different meta-analysis model (fixed effect or random effect) then that was used during the preliminary analysis. 2. By dropping one trial every time. 3. By eliminating trials shorter than two weeks duration. 4. By excluding trials that included trial population with GFR less than 60 mL/min/1.73 m^2^.

In the subgroup and sensitivity analysis above, the rationale for using the benchmark of optimum GFR (60 mL/min/1.73 m^2^) is that SGLT2is is not recommended in T2DM patients with very low GFR.(2) Therefore, the notion is to see if such GFR plays any role in T1DM patients too.

Finally, for the outcomes with statistically significant meta-analytic results, imputation case analysis (ICA) will be done.(29) For ICA, the large dose of SGLTi recipients, and its low dose (or placebo) receivers will be considered as the intervention and comparator arm, respectively. Assuming the event’s occurrence and non-occurrence in all of the missing participants, the ICA-1 and ICA-0 analyses will be conducted, respectively. Moreover, the best- and worst-case scenario will be assessed along with the Gamble-Hollis analysis.(30)

For outcomes that will be statistically significantly different between the compared intervention groups, the evidence will be graded using the GRADE approach.(31)

All analyses will be performed in Stata statistical software version 16 (StataCorp, College Station, Texas, USA).

## DISCUSSION

The chief implication of the prospective review is that it will be the first systematic review that aims to synthesize evidence in this context. It will help physicians, pharma companies, and relevant health authorities to understand how the side effects of SGLTi vary between different doses and with placebo, respectively. Additionally, the proposed study’s findings will allow researchers to compare these with the results of similar research conducted on T2DM patients. Furthermore, this review will also provide the strengths and weaknesses of the existing apposite trials, which may aid future trialists to design better trials.

The proposed review has several strengths. First, since it will be based on double-blinded randomized controlled trials, the highest level of epidemiological evidence, henceforth, the synthesized evidence is likely to be rigorous. Then, since the database search will not be restricted to any date, it will perhaps retrieve an exhaustive list of eligible trials. Finally, the range of proposed sensitivity and imputation analysis will provide an idea of the robustness of the evidence generated from the review.

Despite these strengths, the suggested review is likely to have few weaknesses. At the review level, the inclusion of randomized controlled trials only limits its scope of reviewing studies of other designs like crossover trials or quasi-experimental studies, or good quality observational studies. Besides, limiting the database search to the English language literature only narrows down the scope of reviewing trials published (if any) in other languages. Lastly, since the diagnosis of T1DM and outcome definitions will be accepted as per the trialists, if the trials are extremely heterogenous in this context, the synthesized evidence may be at risk of bias.

## CONCLUSION

The proposed review will compare the safety profile between high and low doses of SGLT inhibitors in insulin-treated T1DM patients. Each of these dosages will be additionally compared with the placebo, respectively.

## Data Availability

not applicable

## CONFLICT OF INTEREST

None declared.

## FUNDING

None received.

## AUTHOR CONTRIBUTIONS

SS^1^ did all work related to preparation of this manuscript.

## ETHICAL APPROVAL

An ethical approval was not required.

## CONSENT FOR PUBLICATION

Not applicable

## AVAILABILITY OF DATA AND MATERIAL

Not applicable

## ACKNOWLEDGEMENTS

Not applicable

## Notes

### Competing Interest Statement

The authors have declared no competing interest.

### Funding Statement

no funding received

